# Clinical Performance and Trends During the First Two Months of Monkeypox Virus PCR Testing at Two United States Reference Labs

**DOI:** 10.1101/2022.09.20.22280169

**Authors:** Nicole A. P. Lieberman, Patrick C. Mathias, Benjamin T. Bradley, Alexander L. Greninger

**Affiliations:** Department of Laboratory Medicine and Pathology, University of Washington School of Medicine, Seattle, WA 98195, USA; Department of Biomedical Informatics and Medical Education, University of Washington School of Medicine, Seattle, WA; Department of Pathology, University of Utah, Salt Lake City, UT; ARUP Laboratories, Salt Lake City, UT; Vaccine and Infectious Disease Division, Fred Hutchinson Cancer Research Center, Seattle, WA

**Author notes:** Co-corresponding authors, Benjamin T. Bradley,; Alexander L. Greninger.

## Abstract

Recently, a sustained human-to-human outbreak of monkeypox virus (MPXV), a member of the *Orthopoxvirus* genus, which includes the etiologic agent of smallpox, has been documented in multiple non-endemic countries including the United States. Prior to June 2022, US testing was limited to public health labs and the Centers for Disease Control and Prevention. Following recognition of the scope of the outbreak, testing for MPXV has expanded into clinical laboratories. Here, we examine epidemiological characteristics, specimen collection practices, and cycle threshold (Ct) values for MPXV PCR tests performed at two reference laboratories.Results from both laboratories support public health data showing a high positivity rate in men (>30%) and those ages 30-49 (25-35%). The overall positivity rate decreased during the study period but remains elevated (∼20%). There was a significant difference in Ct values between laboratories (ARUP 23.86 vs. UW 25.40) and collection method (22.79 for dry swab vs. 24.44 for VTM). These viral load differences likely reflect slight differences in specimen processing. When multiple specimens were collected for a single individual, the overall result concordance rate was greater than 95%, with less than 1.5% of individuals having three or more tests receive a single positive result. As compared to the overall positive cohort, individuals three or more swabs and a single positive result had significantly higher Ct values (22.9 vs 35.0). These results provide an early snapshot of testing in the US during the monkeypox virus outbreak and support restricting the number of swabs collected per individual.

## Introduction

Monkeypox virus (MPXV) is a member of the *Orthopoxvirus* genus that has caused a recent public health emergency of international concern, infecting more than 55,000 persons around in the world and more 21,000 persons in the United States in 2022 as of September 10, 2022. Orthopoxviruses have not spread widely in humans within the United States for approximately 80 years and vaccinia virus vaccination stopped being routine in the United States 50 years ago, leading to substantial gaps in immunity, clinician familiarity with orthopoxvirus infections, and diagnostic testing (1). MPXV PCR diagnostic testing on dry lesion swabs in the United States was initially performed using an FDA authorized orthopoxvirus test run by the Laboratory Response Network (LRN) with confirmatory reflex MPXV-specific testing performed by the CDC (2, 3). In June/July 2022, testing expanded to clinical microbiology laboratories where FDA-authorized and laboratory developed tests (LDTs) are currently available. These LDTs expanded the range of testable specimen types from the initial offering of the FDA-authorized method (dry swabs) to include viral transport media as well as other alternative specimen types. Molecular targets also vary between LDTs and include conserved Orthopoxvirus genus-level targets, probes specific for the West African clade of MPXV, or a multiplexed approach (3, 4). On September 7^th^, 2022, the Secretary of Health and Human Services authorized the emergency use of *in vitro* diagnostics for the detection and/or diagnosis of MPXV infection (5).

As evidenced by the SARS-CoV-2 pandemic, clinical and reference laboratories play a critical role in the response to newly emergent viral diseases in the United States (6). Testing capacities in reference laboratories are often substantially higher than that of the public health system and may provide integrated ordering and reporting of results that can substantially augment detection of viral outbreaks. While qualitative monkeypox virus test results are reported to state health departments and the CDC, aspects of test performance and utility such as cycle threshold (Ct) values or concordance across multiple specimens from the same individual are not readily reported by public health authorities. These data may contain essential information allowing clinical laboratorians to identify emerging viral mutations and steward appropriate testing practices (7, 8). How testing is transitioned from public health laboratories to academic and commercial reference and clinical laboratories at the start of an outbreak remains of considerable interest for American public health and pandemic preparedness. To date, the only published report of MPXV PCR testing data in the United States only described the weekly volumes, positivity rate, and turnaround times from tens of Laboratory Response Network Labs (2). Here, we examined MPXV PCR testing data from more than 10,000 specimens associated with early testing in July and August 2022 from two references laboratories in the western United States, namely ARUP Laboratories (ARUP) in Salt Lake City, UT and the University of Washington Virology Lab (UW) in Seattle, WA.

## Methods

### MPXV PCR testing at UW

UW performs specimen inactivation and lysis in a biosafety cabinet by adding 100µL buffer AL (Qiagen) to 100µL of swab VTM. This combined mixture is used as input to the DNA and Viral NA Small Volume Kit 2.0 (Roche) on the MP96 system for nucleic acid extraction with 100 μL eluate. Each 25μL PCR reaction consisted of the equivalent of 2.71μL water, 11.95μL QuantiTect master mix, 0.1μL each of 100μM forward and reverse primers targeting the MPXV F3L gene (9), 0.05μL of 100μM probe, 0.063μL of an EXO external control primer/probe mix, 0.025μL of UNG enzymes, and 10μL of extracted DNA per reaction. Each reaction was run on ABI 7500 using 2 min 50°C; 15 min 95°C; 45 cycles of 1 min 94°C and 1 min 60°C and positives were called at Ct < 40.

### MPXV PCR testing at ARUP

Presumptive detection of monkeypox virus from clinical samples was performed by a laboratory-developed real-time PCR assay using primer and probes designed by ELITech (Bothell, WA, USA) targeting a conserved region within the polymerase gene common among the Orthopoxvirus family. Samples received on dry swabs were submerged in 500µL of PBS, vortexed for 15 seconds, then allowed to sit at room temperature for 1 hour before proceeding to extraction. For specimens in VTM and dry swabs following resuspension, 200µL of sample was eluted into 80µL on the Chemagic MSMI (PerkinElmer, Waltham, MA, USA) instrument.

Following reaction set-up, amplification was performed on the QuantStudio 12K Flex (ThermoFisher, Waltham, MA, USA) to determine the cycle threshold (Ct). During the study, the the assay was moved to the cobas 6800 (Roche Diagnostics, Indianapolis, IN) platform which includes nucleic acid extraction and amplification. The same primer/probe sequences were used on this platform as the QuantStudio. A method comparison between the two assays demonstrated comparable performance.

### Ethics, Data Analysis, and Statistics

An IRB exemption was granted by the University of Utah IRB under ID#: 00158025. The study was approved by the University of Washington Institutional Review board under STUDY00010205. Deidentified data was analyzed in R v4.0.3. Test results from specimen types other than mucocutaneous lesion swabs were removed by filtering on terms including “serum”, “cerebrospinal”, “breastmilk”, and “blood”. Indeterminate, invalid, and inconclusive results, comprising 0.18% of results, were excluded from analysis. When considering replicate samples submitted for the same individuals, samples collected on different days were not considered part of the same collection event and those individuals were excluded from replicate analysis.

## Results

### Descriptive epidemiology of MPXV PCR testing at ARUP and UW

We first examined the age and sex distributions of individuals tested at each reference lab. When the age distribution of individuals tested at UW was compared to the age distribution in Seattle from the 2020 United States census data (10), it was not significantly different (*p*=0.139, chi squared). Median age of all tested individuals was 33 at both institutions, with an age range of 0-92 (UW) or 0-99 (ARUP). ARUP had a larger proportion of pediatric (age<18 years old) individuals tested for MPXV than did UW (Figure 1A). At UW, the median age of males tested was 34 (range 0-84) and females was 32 (range 0-92). At ARUP, the median age of males tested was 33 (range 0-94) and females was 31 (range 0-99).

**Figure 1.**
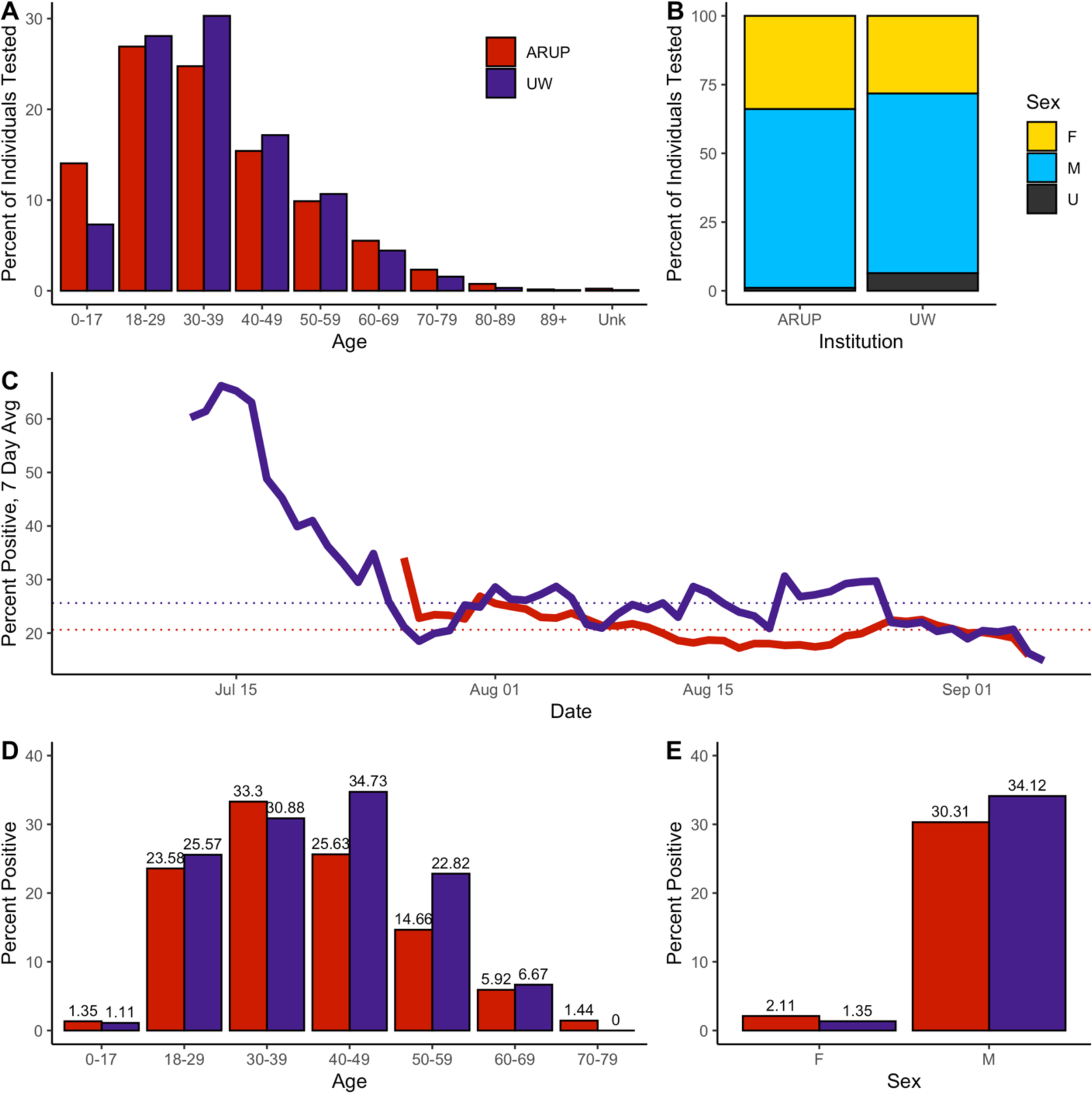
Descriptive epidemiology of individuals tested for MPXV by qPCR at two institutions. A) Age range of individuals tested at ARUP and UW as a percent of all individuals. B) Percent of tests positive for MPXV by age. C) Seven-day rolling average of positive MPXV qPCR test results. D) Sex distribution of individuals tested for MPXV at ARUP and UW. E) Percent of tests positive for MPXV by sex.

At both UW and ARUP, males comprised about 65% of individuals tested for MPXV (Fig1B). Females accounted for a larger share of individuals tested for MPXV than at UW (33.9% vs 28.24%, respectively), while UW had more individuals whose sex was unknown (6.4% vs 1.1%). When individuals with unknown sex were excluded, the distribution was significantly different between the institutions (*p*=0.007, chi squared).

Both ARUP and UW started offering MPXV testing in July of 2022, after which the test positivity rate has decreased (Figure 1C). Driven in part by much higher positivity shortly after the initiation of testing, the cumulative percent positivity has been higher at UW (25.61% vs 20.63%, dotted lines on Figure 1C). The highest test positivity rate at UW was seen in the 40-49 age group (34.83%), while at ARUP, ages 30-39 had the highest rate of positive tests (33.3%) (Fig1D). At both UW and ARUP, males had a much higher test positivity rate than females (34.12% vs 1.35% at UW, and 30.31% vs 2.11% at ARUP) (Figure 1E).

### Comparable Viral Loads in Positive Specimens at ARUP and UW

We also examined factors correlated with and affecting MPXV cycle threshold (Ct) values. Among all positive tests, Ct values at ARUP had an average of 23.86, 1.54 cycles lower than Ct values at UW (mean 25.40), corresponding to approximately 2.9 fold more concentrated viral DNA following extraction assuming similar amplification efficiencies (Figure 2A, *p*=2.9×10^−6^, Welch’s t-test). When Ct values of positive tests were examined by age (Figure 2B), they were consistent across all age groups at UW (*p*=0.722, ANOVA) but displayed variation among ARUP patients (*p*=0.0055, ANOVA). Ct values were the same for males and females at each institution (both: *p*>0.9, Welch’s t-test) (Figure 2C). Among ARUP positive samples, metadata included the type of swab used (dry then submerged in PBS upon arrival at the lab, or in viral transport media (VTM)). Ct values of dry swabs were 1.65 cycles lower on average than swabs in VTM (22.79 vs 24.44, *p*=5.26×10^−8^, Welch’s t-test), or approximately 3.13 times fold more concentrated following extraction of viral DNA (Figure 2D).

**Figure 2.**
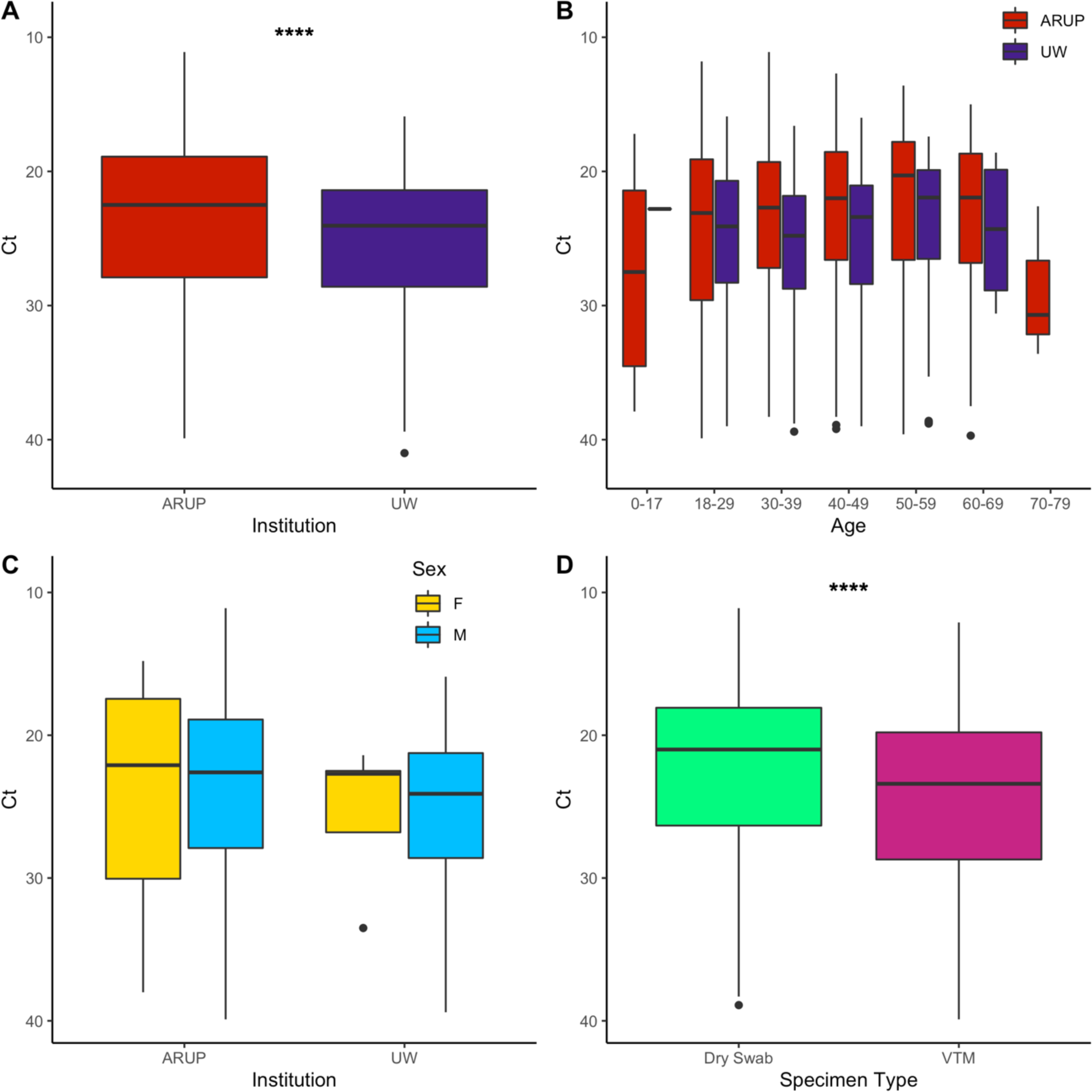
MPXV cycle threshold differences. A) Ct values of positive tests by institution. \*\*\*\**p*<0.0001, t-test. B) Ct values of positive tests by age. C) Ct values of positive tests by sex. D) Ct values of positive tests by swab type. \*\*\*\**p*<0.0001, t-test.

### High Concordance of Multiple Swabs Sent from the Same Individual

We have observed wide variation in the number of samples submitted for testing for each individual. While the majority of individuals had only a single sample submitted for testing, more than 31% of ARUP patients and more than 10% of UW patients had two or more specimens collected at the same time (Figure 3C, data was filtered to exclude tests not collected on the same day). To explore the value of testing multiple swabs from the same patient, we examined test result concordance, defined as all results returning positive or all results returning negative. Among individuals with two, three, or four replicate samples submitted for testing, concordance exceeded 95% (Figure 3B). Notably, among individuals that received three or four samples tested, only 1.3% and 2.4% had only a single positive test (Figure 3C).

**Figure 3.**
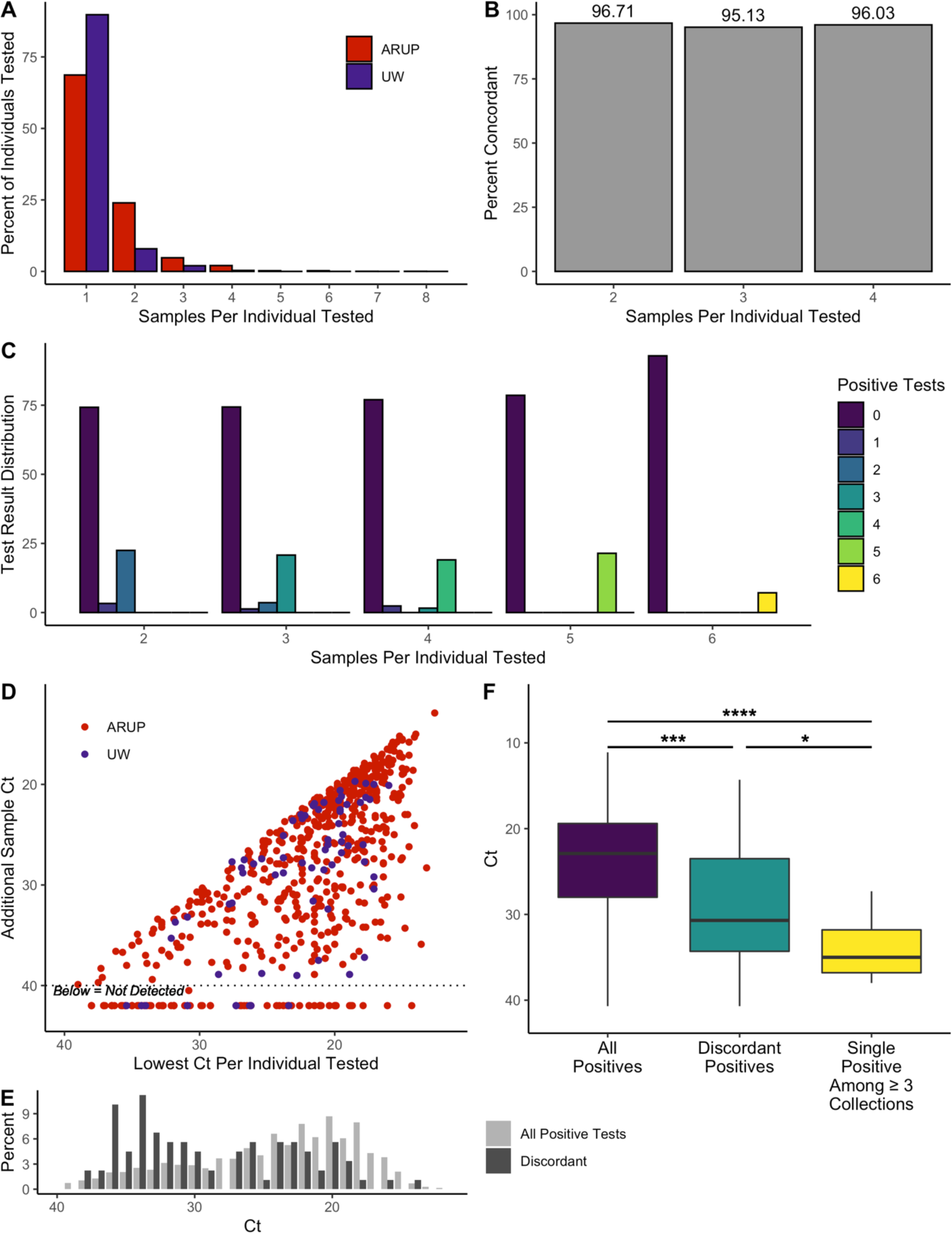
Examination of the yield and discordant results for individuals with multiple tests taken on the same collection date. A) Percent of all individuals by number of replicate tests. B) Among replicate tests, percent of individuals receiving all positive or all negative (concordant) test results. C) For each group of n replicate samples per individuals, the percent of individuals receiving 0, 1, or more positive tests. D) Comparison of Ct values for individuals that had two or more replicate specimens collected. E) Distribution of Ct values for individuals testing positive with discordant results (at least one negative) versus of all positive tests. F) Median and interquartile range of Ct values of positive results among all samples, among positive samples with at least one paired negative sample (discordant positives), and among samples that were the single positive test result when 2 or more additional replicate samples tested negative. * *p*<0.05, *** *p*<0.001, **** *p*<0.0001.

We speculated that individuals with discordant results may have lower viral burden and correspondingly higher Ct values. Figure 3D shows a scatter plot of paired Ct values among individuals with at least one positive test result. The lowest Ct is plotted on the x axis and any matched Ct value is plotted on the y axis. A strong positive relationship was observed (Pearson coefficient = 0.695), indicating that in most cases a low Ct value in one sample is likely to be repeated in a paired sample. However, when only the Ct values from individuals with discordant results are plotted, a skew toward higher Ct values relative to all positive samples can be seen (Figure 3E, dark grey vs light grey). Among all positive tests, only 18.4% of Ct values are higher than 30, while among discordant tests, 51.69% of Ct values are higher than 30. This is made more clear in Figure 3F, which shows the median Ct for all positive tests (22.9), positive tests from individuals with discordant results (30.7), and the single positive test from individuals with three or four replicate samples (35.0) (*p*<2e^-16^, ANOVA).

## Discussion

Here, we detail approximately eight weeks of MPXV PCR testing data from July and August 2022 from two large reference laboratory laboratories. Our report substantially adds to the data available on monkeypox testing in the United States, as the main prior testing report from the CDC detailed testing of 2,009 specimens over a six-week period from late May to June 2022 (2). Overall, testing was enriched in the population at greatest risk for monkeypox (males ages 18-40). We found a decline in positivity rate over time as more specimens were submitted, though positivity rates persisted at relatively high levels (∼20%). We found a substantial difference in positivity rates between specimens submitted from males over that of females, consistent with prior reports of the vast majority of infections occurring in men who have sex with men (11).

The viral load of positive specimens was comparable between ARUP and UW, with a trend toward higher viral loads at ARUP. We hypothesize that viral load differences are due to extraction protocols, as UW extracts half the material of ARUP in order to add lysis buffer in a biosafety cabinet prior to use of extraction instrumentation. In addition, ARUP uses the Chemagic nucleic acid extractor which was previously associated with some of the better analytical sensitivities for COVID-19 PCR (12). Unsurprisingly, dry swabs were associated with stronger viral loads than swabs in VTM since they are resuspended 6-fold less liquid and thus are more concentrated. Nonetheless, the workflow limitations to dry swab testing restrict the ability to test these specimens in a high-volume reference laboratory.

As CDC recommendations encourage collecting two swabs, we specifically examined our testing data to understand the utility of submitting multiple lesion swabs for detection of MPXV. Skin lesions have already shown the highest sensitivity (13) and submitting multiple specimens in separate tubes increases the labor demands on clinic and laboratory staff and may substantially increase costs associated with sending MPXV testing. While the vast majority of individuals had only one specimen submitted, approximately one-third of individuals tested at ARUP had multiple specimens submitted, suggesting a considerable portion of MPXV PCR testing is associated with this use case. Testing results for multiple specimens were highly concordant and we found 1.5% of individuals with three or more swab submitted had only one swab test positive. Discordant positives were also associated with substantially lower viral loads than the total positive cohort. These discordant results may be explained by one of two major causes: First, these samples may have contained a low viral load with stochasticity near the limit of detection affecting sensitivity. Alternatively, discordant samples may be false positives resulting from cross-contamination by an adjacent high viral load specimen. On August 23^rd^, the CDC issued a Lab Advisory warning of false positive risk and recommended repeat testing of samples with Cts greater than 34 (14). However, a specimen which repeated as negative does not rule out stochastic effects for samples near the limit of detection., suggesting sampling stochasticity near the limit of detection could be the main driver of sensitivity. Overall, our data suggest current CDC recommendations of sampling at least two lesions per patient provides sufficient diagnostic performance for the vast majority of patients (15). With the availability of VTM testing now, we believe the most practical method for specimen collection is to combine swabs into a single tube of VTM when collected from the same source in order to reduce overall testing costs.

Limitations of this work include its descriptive nature, restriction to lesion swabs, and use of reference laboratory data with sparse clinical metadata. We did not specifically consider diagnostic yield in non-standard specimen types as data for these specimens was very sparse. Furthermore, determining the utility of specific specimen types is best performed in the context of prospective studies, although retrospective, descriptive examinations of clinical testing data can generate hypotheses for how diagnostic testing is being used in the real world. For those few individuals who had discordant results associated with low viral loads, we are unable to draw on additional or orthogonal testing data or electronic health record data to adjudicate the likelihood of a false positive (16). Nor were we able to determine if any asymptomatic individuals tested positive, as has been recently reported (17). The availability of MPXV serology would help adjudicate these cases, however currently no authorized commercial MPXV serological assays nor fully MPXV-specific assays exist. It is likely more work is needed on transitioning accurate MPXV serology testing from CDC to academic and clinical reference laboratories in a similar manner as the MPXV PCR testing described here.

## Data Availability

All data produced in the present study are available upon reasonable request to the authors

## Notes

### Competing Interest Statement

ALG reports contract testing from Abbott, Cepheid, Novavax, Pfizer, Janssen and Hologic and research support from Gilead and Merck, outside of the described work.

### Funding Statement

This study did not receive any funding

### Author Declarations

An IRB exemption was granted by the University of Utah IRB under ID#: 00158025. The study was approved by the University of Washington Institutional Review board under STUDY00010205

